# A 20-year experience with initial and repeated electro- and echocardiographic screening in youth, athletes, and adults revealing high number of abnormalities

**DOI:** 10.1101/2024.05.07.24307029

**Authors:** Sharon Bates, Mohammad Reza Movahed

## Abstract

**Background:** To further analyze the impacts, findings, and modalities of multiple cardiac screenings to answer the question – are multiple screens necessary in youth?

**Methods:** Over 20 years, the Anthony Bates Foundation (ABF) has offered free and low-cost cardiac screenings to youth and their families nationwide. The volunteer force has provided blood pressure, and ultrasound tests to participants throughout the 20 years. After year 7, electrocardiograms were added to the screening.

**Results:** Over the 20 years, ABF abnormal findings held steady between 10 - 13%, with Potential Life-Threatening (PLT) findings at 2.5%. The participants that have experienced multiple screening tests on average would repeat within 2.5 years, have abnormal findings at 31.84%, PLT at 11.43%, and total echocardiography-related abnormal findings at 16.82%. The variance between male and female attendance by age is also noted during the review of ABF repeat screened data. Male attendance was at 59.65% and female 40.35%. The abnormality rate of males for the first visit was 10.9% followed by the second visit of 18.80%. The abnormality rate of females for the first visit was 12.22% followed by the second visit of 17.09%. A detailed analysis of abnormal findings is presented in this manuscript.

**Conclusions:** Cardiac screening involving multiple repeated screenings appears to be effective in detecting increasing numbers of abnormal findings that can be lifesaving.

## Introduction

The tragedy of a young life lost to underlying and often unsuspected heart disease (1) became a reality for Sharon Bates. July 2000, her son, Anthony Bates died from undiagnosed Hypertrophic Cardiomyopathy (HCM) while a college football player. His story and the reason for starting screening is described in a book in detail (2) On average, every 3 days in the United States a competitive athlete experiences a Sudden Cardiac Death (3). She started providing cardiac screenings for youth and families and then formed the Anthony Bates Foundation (ABF) to build a legacy for her son. Learning over the years of screening, additional studies suggested proponents of ECG screening in the United States suggest that it can be effective, feasible, and cost-effective (4). Passionate, emotional, and compelling arguments have been advanced for the implementation of national screening programs such as the Italian approach (3). In a recent report from the Institute of Medicine, of the estimates of sudden cardiac arrest deaths (395,000 annually), approximately 12,500 are children (5). That calculates to 250,000 dead children since Anthony died from undiagnosed HCM at age 20. The emergency of this public health crisis is well overdue. The goal of this study was to evaluate the effect of repeat screening on the number of abnormal findings.

## Method

The cardiac screenings provided by ABF include a two-page questionnaire with health history and cardiac questions, a blood pressure check, a 12 lead electrocardiogram (ECG/EKG), and a limited ultrasound with measurements on the septal wall, left ventricle size, posterior wall, and the left ventricle outflow tract. The ECG tests were added for all participants in 2007. During the large, 6-hour-long, ABF heart-screening events an average of 150 tests were performed per day. The involvement in administering the event and providing such tests are done by community volunteers (junior high, high school students, and adults), and community medical volunteers (nurses, nursing students, emergency medical technicians, cardiac ultrasound technicians, cardiac ultrasound students, and cardiologists.) Small events were offered in-office to an average of 15 people and a paid cardiac ultrasound technician assisted in providing ECG and ultrasound tests. All screening results are reviewed by a volunteer cardiologist. The database contained the questionnaire answers, the heart-screening test results, and comments by the reading cardiologist. Over a 20 year period from 2000 to 2020, ABF has offered free and low-cost cardiac screenings to communities in Arizona, Kansas, Colorado, Washington, and New York.

## Statistical Analysis

A comparison of the abnormal results from the overall 20-year ABF heart-screening database is calculated from person to abnormal findings. The simple division of abnormal findings (2075) into the total screened (15,175) multiplied by 100 creates the abnormal percentage (13.7%). If we remove the multiple heart-screened patients/participants (446), there could be a smaller abnormal percentage. Experience indicates that when there is disagreement among experts, there is an abundance of reliable data (Link, 2012). More research would be necessary to determine the suitable statistics of the abnormal findings. Using that same simple division equation above for the repeat heart-screens using abnormal findings for repeat attendance (142) divided by the patient attendance (446), not the number of multiple screening records (952), and then multiplied by 100 will bring the percent of abnormal findings for the repeat heart-screens (31.84%).

The abnormal percentage is broken down into the initial/first heart screening and second screening for the 446 attendees. There were 51 abnormal findings during the initial screening, divided by the 446 attendees, and multiplied by 100 to get the abnormal findings for the initial heart-screening tests at 11.43%. The initial heart-screening percentage of 11.43% is 2.27% less than the overall ABF heart-screen database at 13.7%.

To analyze the abnormal results findings there must be awareness that a patient/participant can and often does have more than one finding. For example, a patient/participant could have high blood pressure and an ECG abnormality or any combination of the abnormal finding categories. Therefore, a comparison of the number of abnormal findings is not compared to the number of attendees. The analysis of the abnormal findings shown in the figures is a comparison by percent of the total of abnormal findings. This comparison was done for the overall ABF heart-screening database and for the return patient/participants in the initial heart-screening database and the second heart-screening database.

Careful analysis was also done by age and gender for the return patient/participants for the initial heart-screening tests and the second heart-screening tests. The percentage of abnormal heart-screening tests rose from the initial tests of 11.43% to 17.49% for the second test. Also noted was the increase of abnormal heart-screening test results in youth under the age of 19 from the initial heart-screening (10.26%) to the second heart-screening (12.33%). Combining all patients/participants heart-screened raises the percentage to 31.84%, well above at an increase of 18.14% to the overall ABF heart-screened of 13.70%.

A brief comparison of socio-economic was also included in the statistical abnormal heart-screening finding of the repeat patients/participants. For the repeat patients/participants, 62% of abnormal heart-screening findings were in the up to $20k (poverty) level. Keeping in mind the students and youth in this category may not have income, and not include their parents’ or guardians’ income.

## Results

The data from these 105 large heart screening events and 94 small heart screening events has captured 15,175 records in the ABF database. The cardiac screening tests revealed an abnormality rate of 10% increasing to 13% over the years. ABF has evaluated the heart-screening result tests to be possible-life threatening in 2.5% of overall abnormal results. Of the ABF heart screening participants, 446 people have rescreened their hearts during multiple events. The ABF database contains 954 records of the repeat participants. The return participant on average returned to ABF within 2.5 years. 78.70% of the return participants were below the age of 26 (college and youth.) There were 59.65% of return participants male. The abnormality rate for males grew from 10.9% on their first heart screening to 18.8% on their second heart screening. Whereas, the abnormality rate for females went from 12.22% on the first heart screening to 17.09% on the second heart screening.

Limited reviews of the socio-economics of the people with abnormal findings during the repeat heart screening revealed 62% were in the up to $20k (poverty) level and 66.7% of the possibly life-threatening (PLT) findings were in the same $20k (poverty) level. The overall screening totals of 20 years’ worth of data have shown a much lower 17% of abnormal findings in the up to $20k (poverty) level.

The repeat screening tests revealed a higher-than-expected abnormality rate of 31.84% overall. The initial tests showed an ABF lower than average rate of 11.43%, but the second heart screening had a higher rate of 17.49%. Figure 1 and 2. Echocardiography found over half the abnormal findings, 75 out of 142, during the initial and second heart-screens. Follow-ups of abnormal findings were initiated by ABF and volunteer nursing students in 2014. Patient interaction and confirmation of findings were limited by 1 out of 15 callbacks. Of the interactions and surveys taken the feedback was overwhelmingly positive.

**Figure 1:**
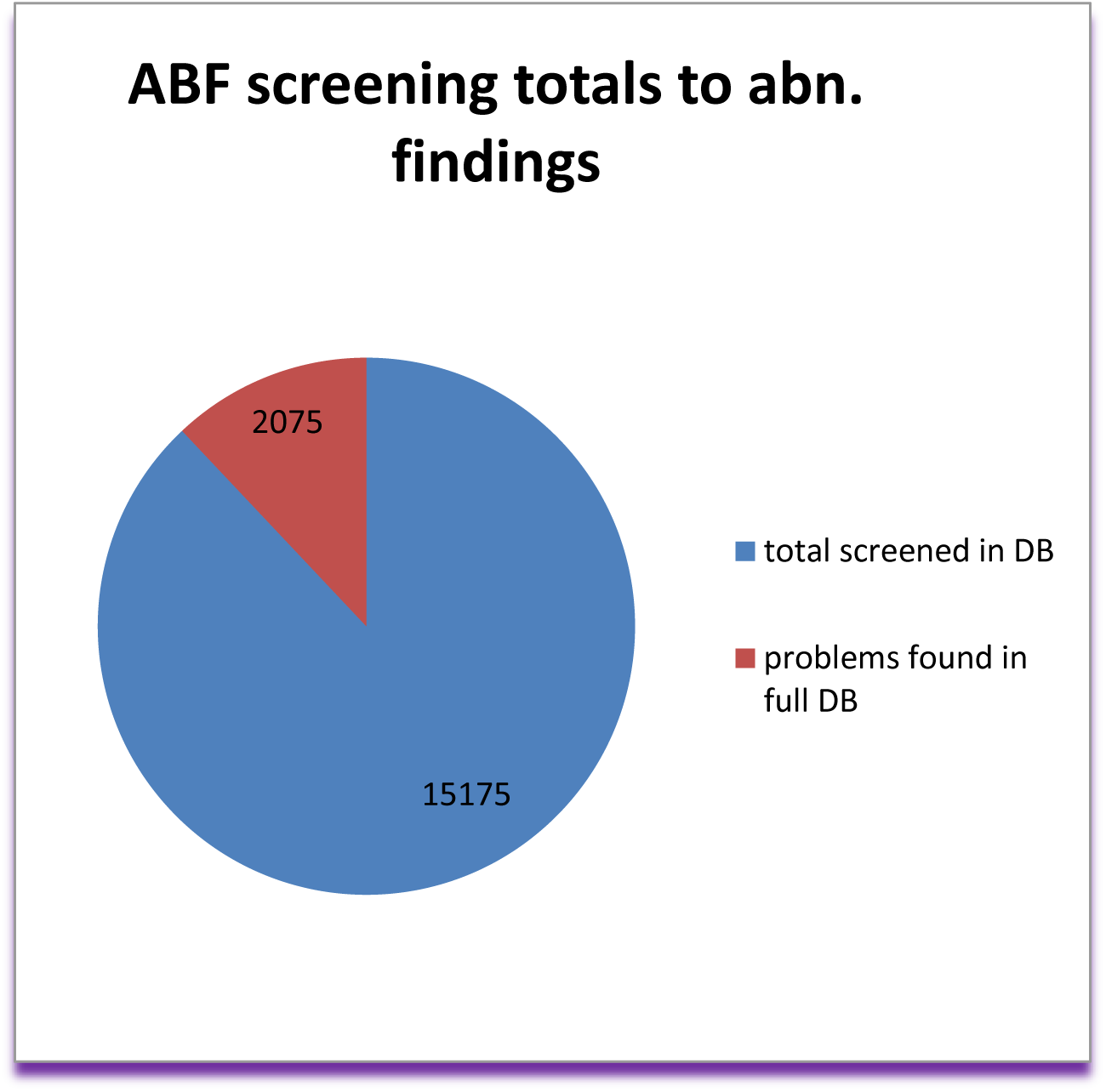
ABF initial Screening with Abnormal findings

**Figure 2:**
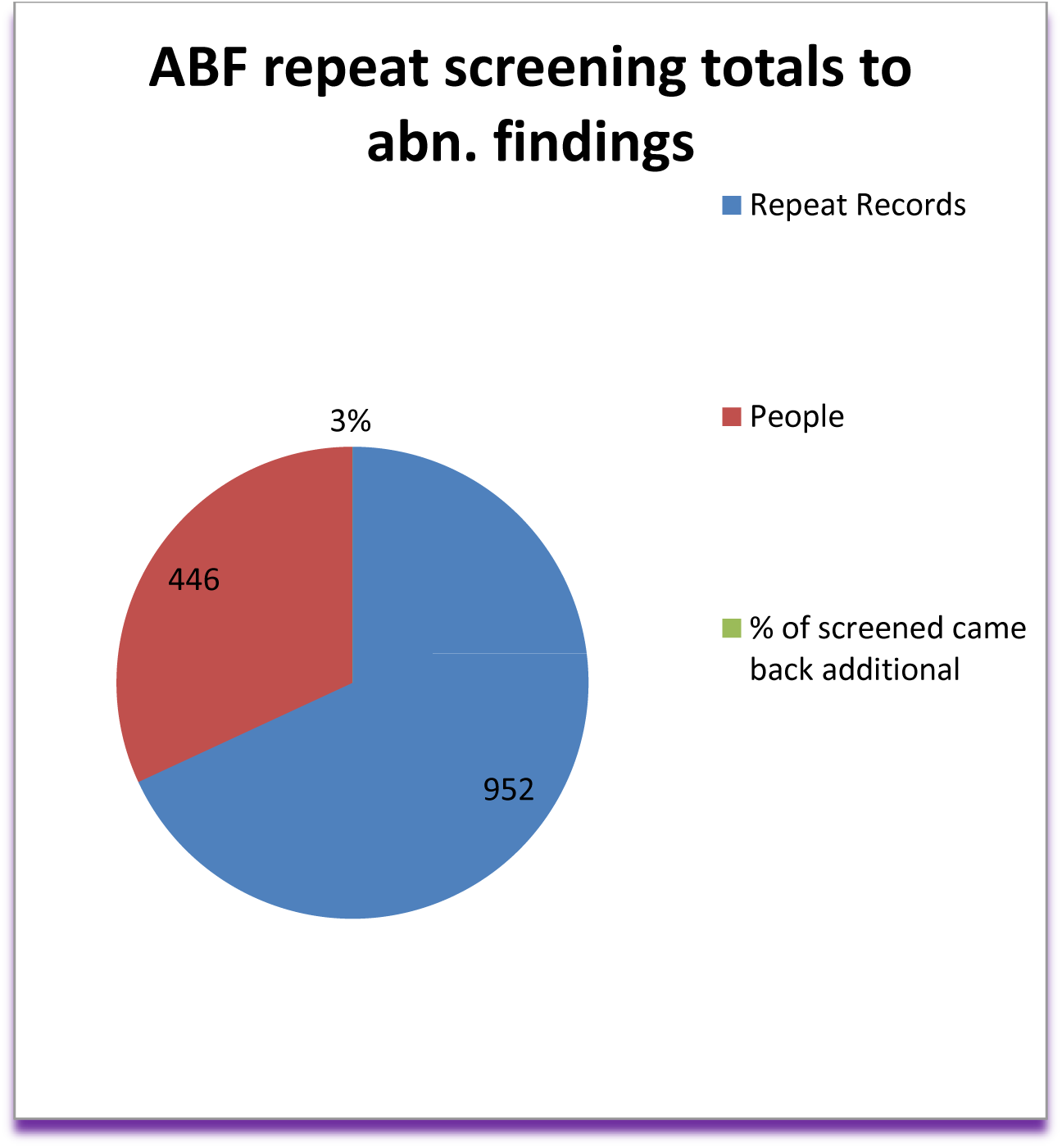
ABF Repeat screening and abnormal findings

The abnormal findings for the total 2075 participants/patients within the ABF database are represented in Figure 3 by a percentage. Several of the participants/patients have multiple findings. ABF shows abnormal results as follows: blood pressure (39.57%), ECG irregularities (28.34%), valve problems to include MV and other valve issues (11.95%), chamber abnormalities including LAE, LVH, and RVH (13.35%), follow-up on symptoms (7.86%), VSD, ASD, PDA – shunts (2.22%), possible Marfans Syndrome (1.25%), and BAV (0.24%). Nearly 30% of the ABF Overall Findings are found using echocardiogram tests. Therefore, ABF or other screening programs would miss nearly 30% of the abnormal findings without echocardiogram screening tests.

**Figure 3:**
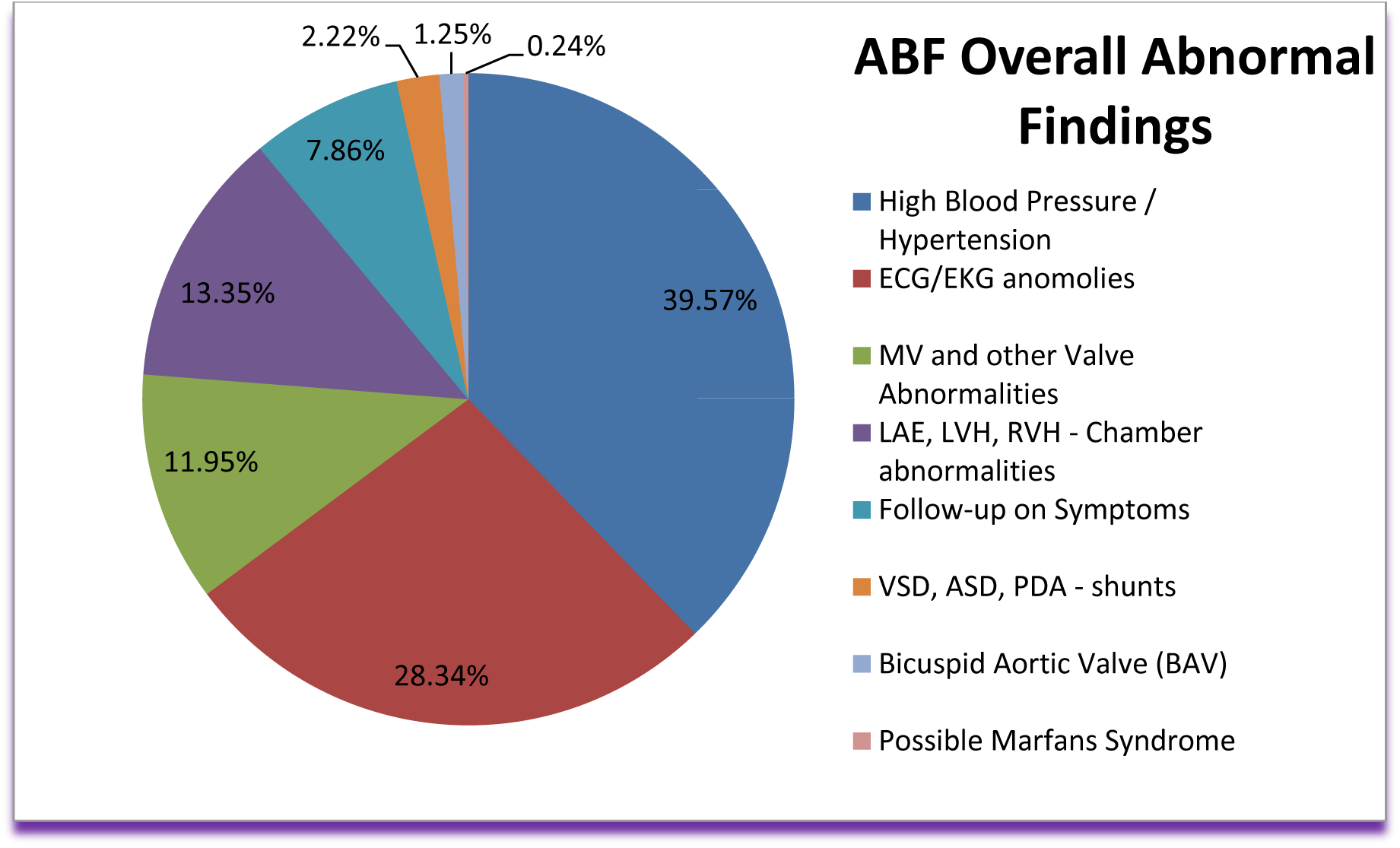
ABF Overall Abnormal Findings in details

The abnormal findings for the multiple or repeat screenings can be seen in Figure 4 represented as a percentage. Several of the participants/patients have multiple findings. ABF shows abnormal results as follows: blood pressure (13.23%), ECG irregularities (8.97%), chamber abnormalities including LAE, LVH, and RVH (8.07%), valve problems to include MV and other valve issues (7.17%), follow-up on symptoms (1.35%), VSD, ASD, PDA – shunts (1.35%), possible Marfans Syndrome (0.45%), and BAV (0.22%). The number and type of abnormal findings of the initial screen tests are represented in Figure 5. The abnormal findings are as follows: VSD, ASD, PDA – shunts (1); Follow-up on symptoms (5); MV and other valve abnormalities (15); LAE, LVH, RVH – chamber abnormalities (18); ECG irregularities (10); and blood pressure issues (16).

**Figure 4:**
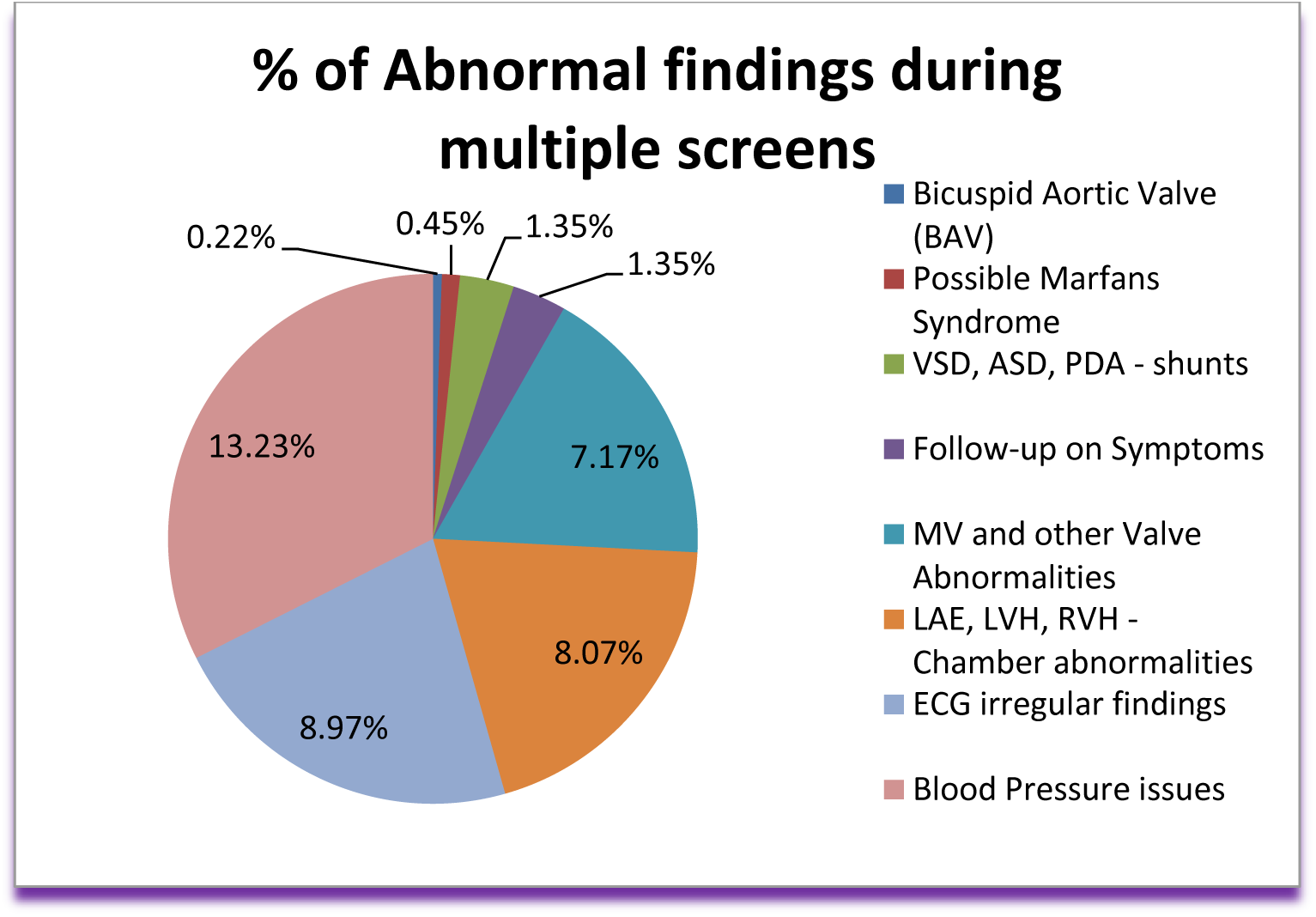
Percentage of Abnormal findings during multiple screening in details

**Figure 5:**
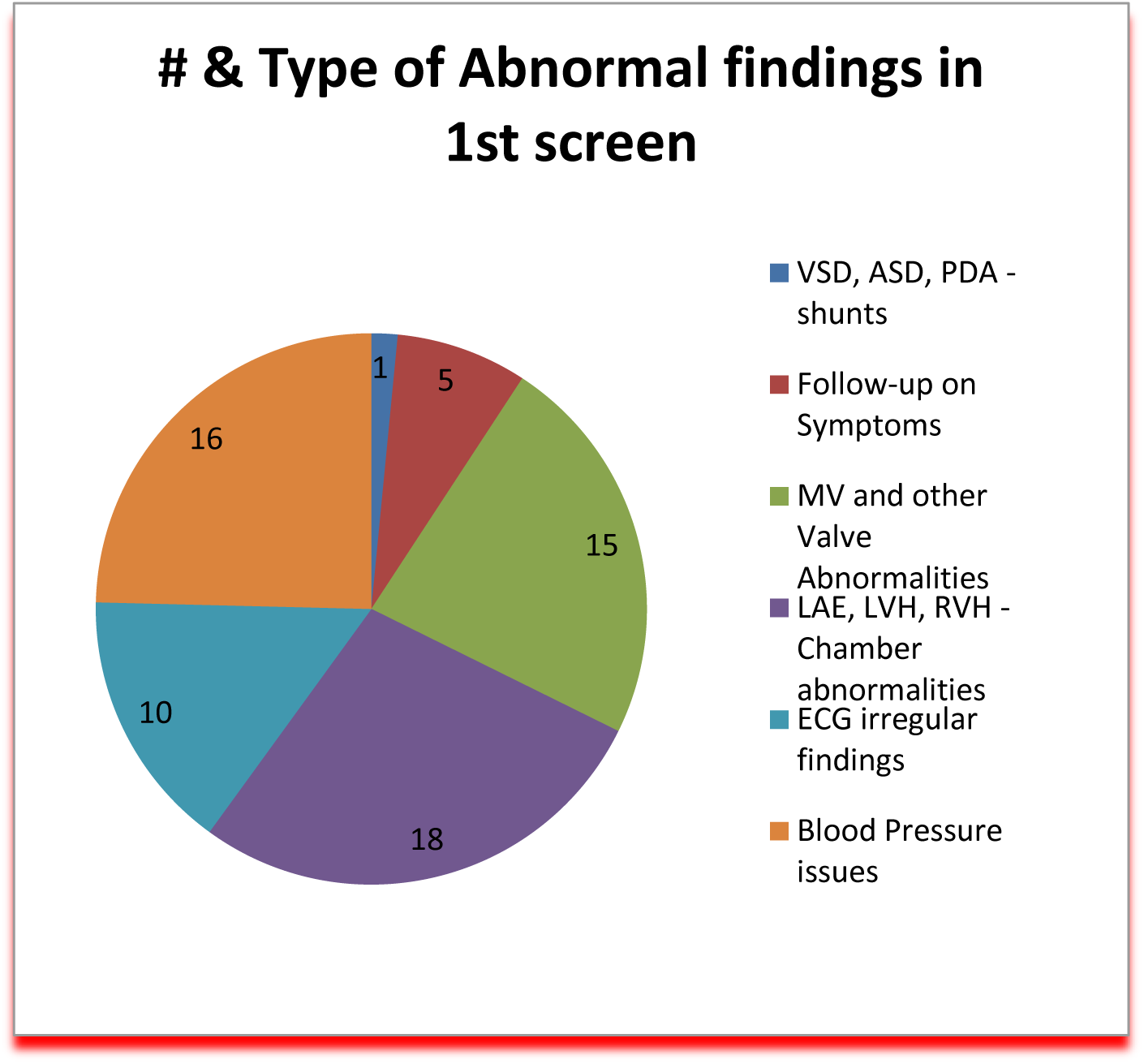
Type of Abnormal findings in 1st screen

The number and type of abnormal findings of the second screen tests are represented in Figure 6. The abnormal findings are as follows: BAV (1); possible Marfan Syndrome (2); VSD, ASD, PDA shunts (4); Follow-up on symptoms (1); MV and other valve abnormalities (11); LAE, LVH, RVH – chamber abnormalities (22); ECG irregularities (28); and blood pressure issues (33). Figure 7 represents the comparison of the percent of abnormal findings to the total patients/participants in three screenings evaluated in this paper. The overall 20-year ABF database of all attendees shows an average abnormality rate of 13.7%. The middle set of data represents the repeat patients/participants of the initial screen at an abnormality rate of 11.43%. The last set of data represents the repeat patients/participants of their second screen with an abnormality rate of 17.49%. Figure 8 represents the combined abnormal findings of the repeat patients/participants who attended ABF screenings. There were 142 people with abnormal findings, 51 had possible life-threatening (PLT) findings, and 77 of the findings were found with echocardiogram tests. The data in Figure 9 represents the abnormal findings on the repeat screen patient/participants during their initial screen. In these initial screenings, there were 51 patients/participants with abnormal findings, 14 of those were possibly life-threatening (PLT), and 29 of the abnormal findings were found through echocardiogram tests. The abnormal findings of the patient/participants of the second screen tests are represented in Figure 10. There were 78 people found with abnormal findings, 32 were possibly life-threatening (PLT) and 23 were found with echocardiogram tests.

**Figure 6:**
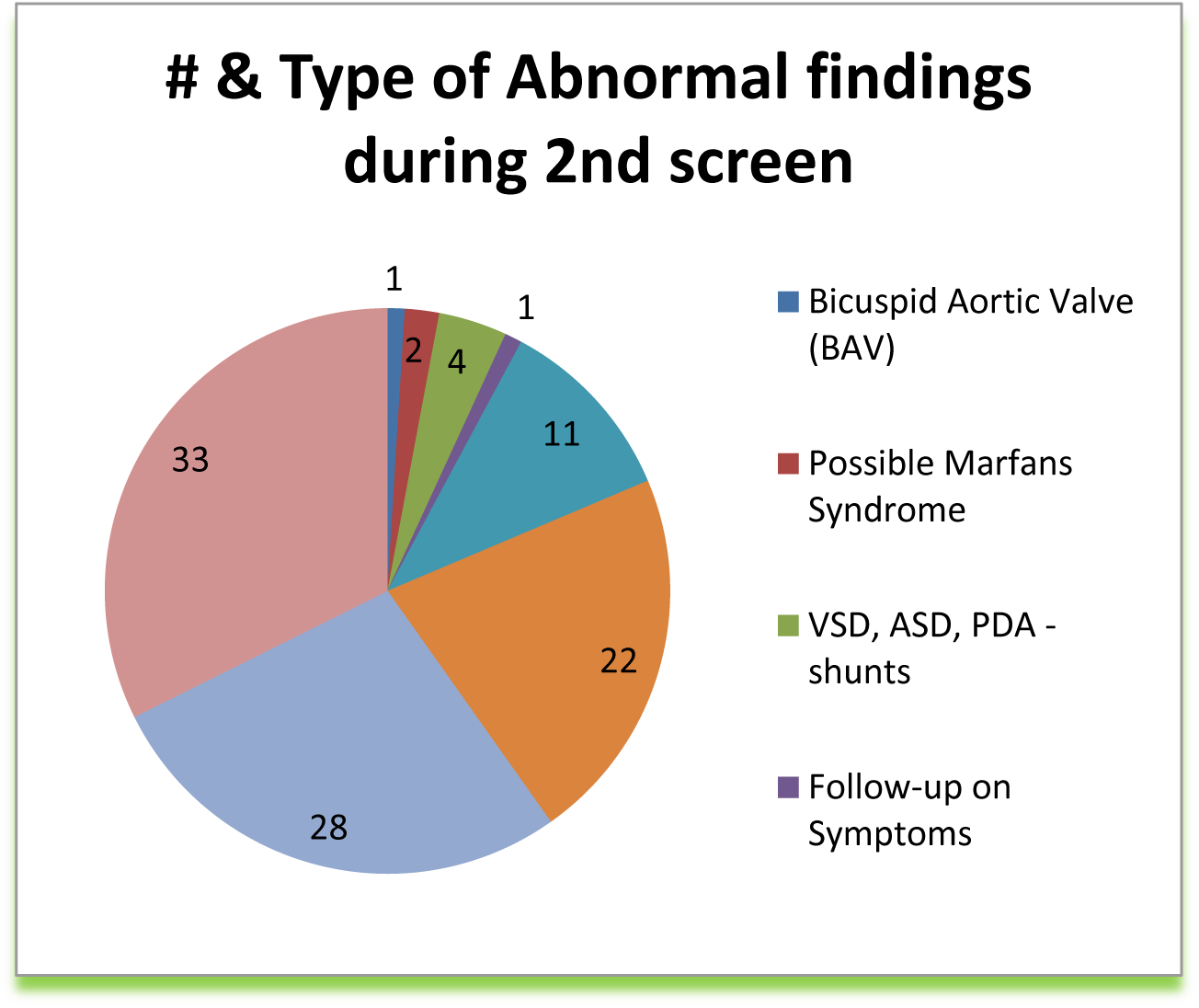
Type of abnormal findings during 2nd screen

**Figure 7:**
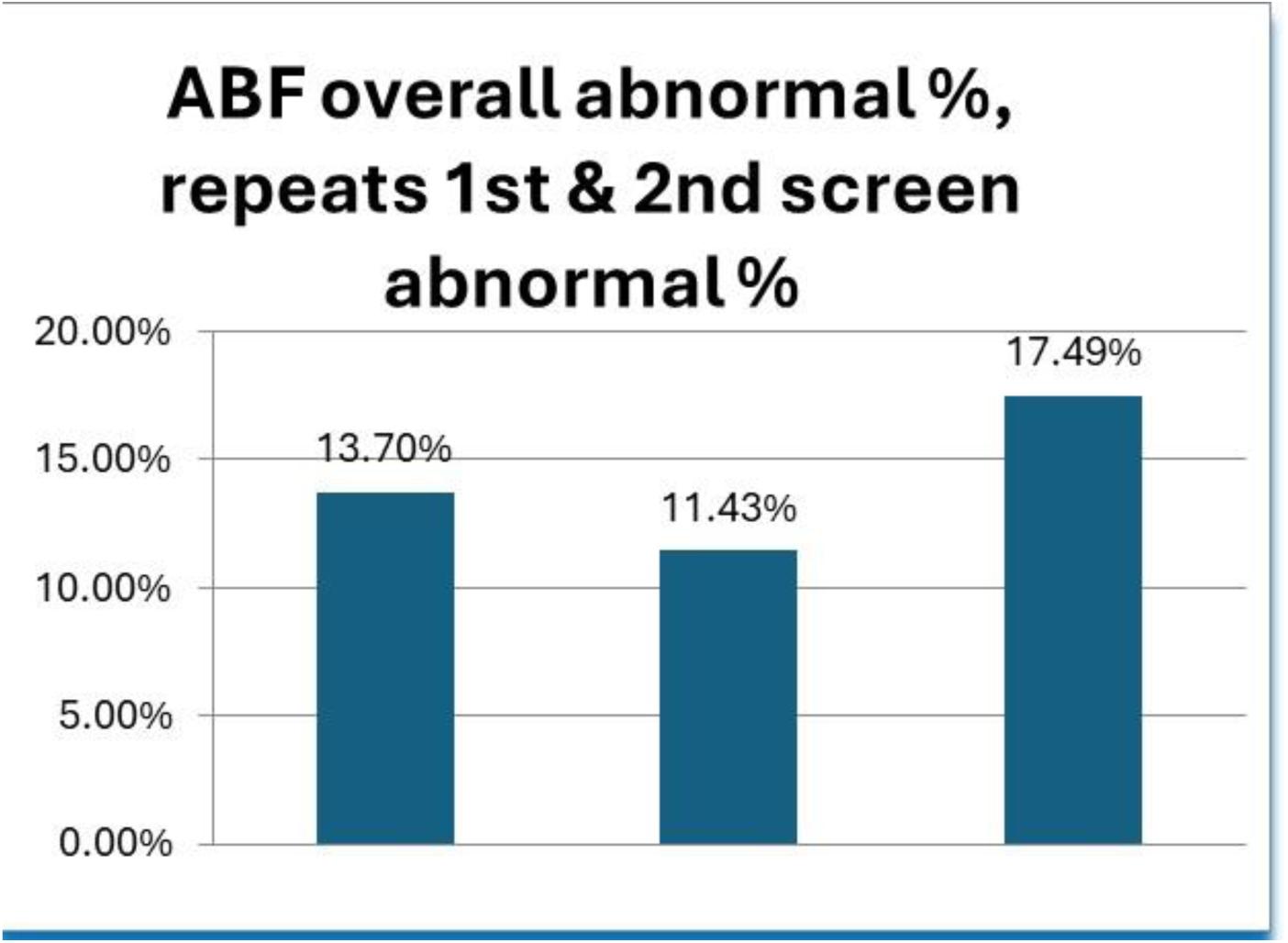
ABF Abnormal % of Overall screens, then repeats 1st & 2nd Screens

**Figure 8:**
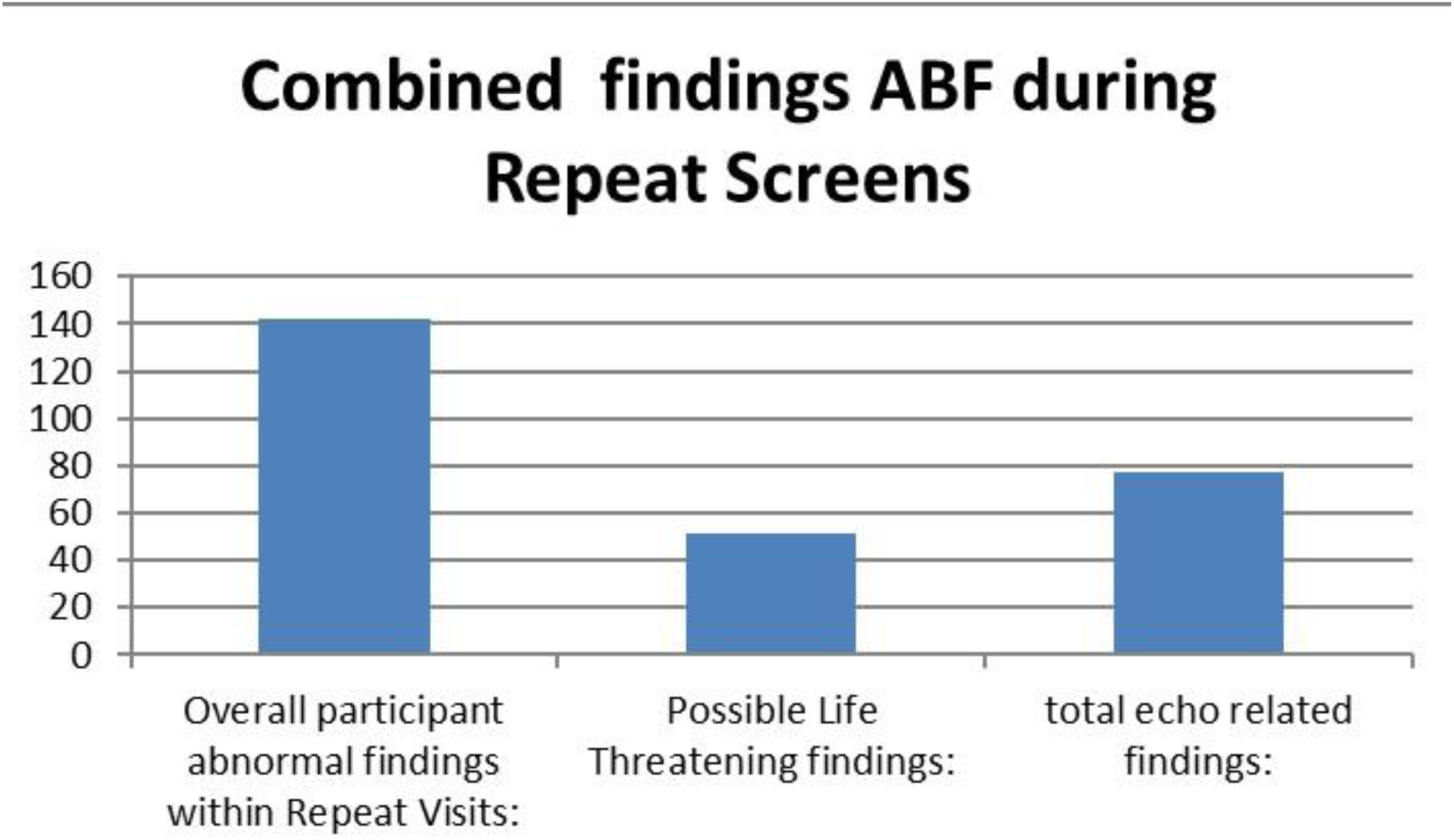
ABF Repeat Screens abnormality compared to PLT and Echo findings

**Figure 9:**
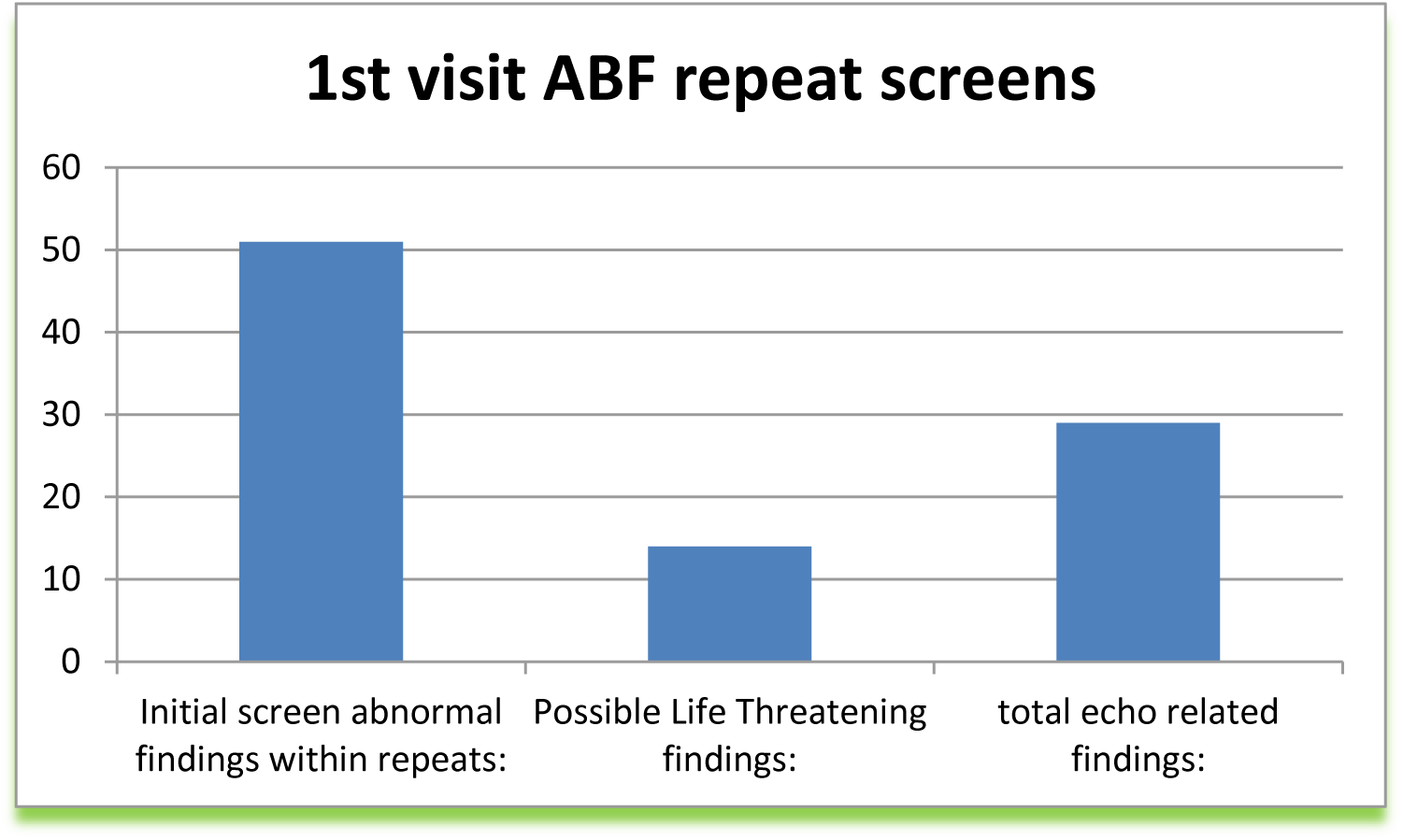
Abnormal findings of the 1st screen

**Figure 10:**
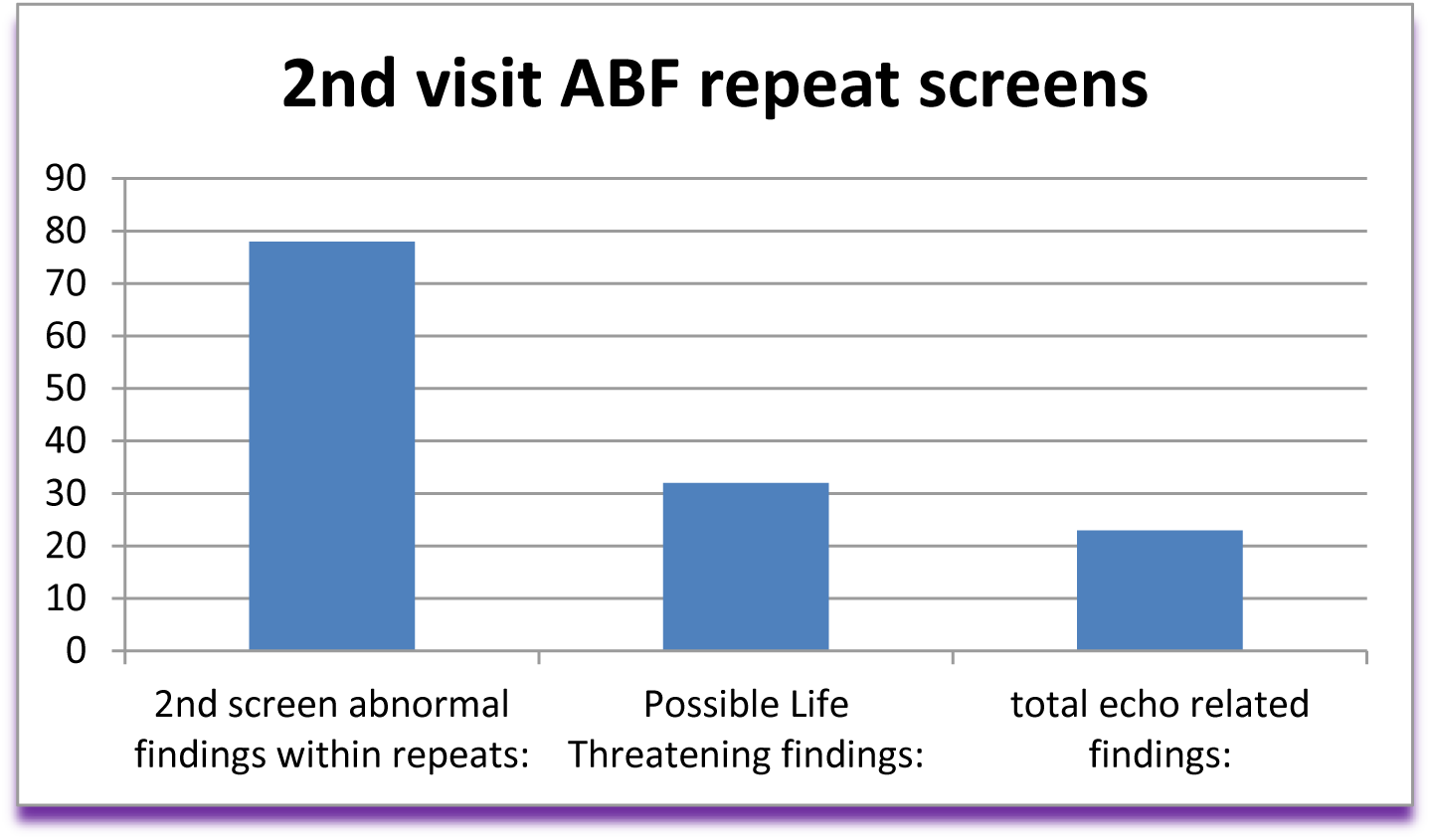
Abnormal findings of the 2nd screen

Table 1 is the representation of the gender comparison for the 446 repeat patients/participants. Table 2 is the representation of the gender and age comparison to the abnormal findings with numbers and percentages of each gender/age category. The purpose of this breakdown is to recognize the high youth abnormality rates compared to older adults. Table 3 represents a rudimentary analysis of socioeconomic categories for the repeat patients/participants who had abnormal heart-screening findings.

**Table 1:** Repeat Screens by Gender.

|  |  |  |  |
| --- | --- | --- | --- |
| Repeat |  |  |  |
| Screens | 59.64% | 266 | Male |
| Gender | 40.36% | <u>180</u> | Female |
|  |  | 446 |  |

**Table 2:** Repeat pts/participants with gender and age comparison to abnormal findings.

| <u>First Screening</u> |  | <u>Males</u><br><u>w/findings</u><br><u>1st Visit</u> |  | <u>PLT</u><br><u>Findings</u> |  |
| --- | --- | --- | --- | --- | --- |
| 194 | Males below 19 Y/O | 18 | 9.27% | 1 | 0.51% |
| 44 | Males Between 19 & 25 Y/O | 6 | 13.60% | 3 | 6.80% |
| 24 | Males Between 26 & 56 Y/O | 5 | 20.83% | 3 | 12.50% |
| 4 | Males Older Than 56 Y/O | 0 | 0.00% | 0 | 0.00% |
| 266 |  | 29 | 10.90% | 7 | 2.63% |

| <u>First Screening</u> |  | <u>Females</u><br><u>w/findings</u><br><u>1st Visit</u> |  | <u>PLT</u><br><u>Findings</u> |  |
| --- | --- | --- | --- | --- | --- |
| 72 | Females below 19 Y/O | 9 | 12.50% | 3 | 4.16% |
| 41 | Females Between 19 & 25 Y/O | 3 | 7.31 | 0 | 0.00% |
| 49 | Females Between 26 & 56 Y/O | 5 | 10.20% | 1 | 2.04% |
| 18 | Females Older Than 56 Y/O | 5 | 2.77% | 2 | 1.11% |
| 180 |  | 22 | 12.22% | 6 | 3.33% |
|  | 11.43% | 51 |  |  |  |

| <u>Second Screening</u> |  | <u>Males</u><br><u>w/findings</u><br><u>2nd Visit</u> |  | <u>PLT</u><br><u>Findings</u> |  |
| --- | --- | --- | --- | --- | --- |
| 167 | Males below 19 Y/O | 13 | 7.78% | 5 | 2.99% |
| 68 | Males Between 19 & 25 Y/O | 23 | 33.82% | 11 | 16.18% |
| 23 | Males Between 26 & 56 Y/O | 12 | 52.17% | 5 | 21.74% |
| 8 | Males Older Than 56 Y/O | 2 | 25.00% | 0 | 0.00% |
| 266 |  | 50 | 18.80% | 21 | 7.89% |

| <u>Second Screening</u> |  | <u>Problems</u><br><u>Found</u><br><u>2nd Visit</u> |  | <u>PLT</u><br><u>Findings</u> |  |
| --- | --- | --- | --- | --- | --- |
| 48 | Females below 19 Y/O | 6 | 12.50% | 2 | 4.17% |
| 71 | Females Between 19 & 25 Y/O | 13 | 18.31% | 6 | 8.45% |
| 45 | Females Between 26 & 56 Y/O | 5 | 11.11% | 1 | 2.22% |
| 16 | Females Older Than 56 Y/O | 4 | 25.00% | 2 | 12.50% |
| 180 |  | 28 | 17.09% | 11 | 7.04% |
|  | 17.49% | 78 |  |  |  |

**Table 3:** Socio-economic breakdown for pts/participants with abnormal findings.

| | | Up to<br>\$20k | \$20K to<br>\$50K | \$50K to<br>\$75K | \$75K to<br>\$100K | Over<br>\$100K | |
| --- | --- | --- | --- | --- | --- | --- | --- |
| Abnormals | 142 | 88 | 18 | 12 | 7 | 17 |  |
|  | 100.00% | 62% | 12.60% | 8.40% | 4.90% | 11.90% |  |
| PLT | 51 | Up to<br>\$20k | \$20K to<br>\$50K | \$50K to<br>\$75K | \$75K to<br>\$100K | Over<br>\$100K | |
|  |  | 34 | 7 | 5 | 1 | 4 |  |
| % of PLT | 100.00% | 66.7% | 13.7% | 9.8% | 2.0% | 7.8% |  |
| % of<br>Abnormals | 34.20% | 22.3% | 4.9% | 3.5% | 0.7% | 2.8% |  |

## Discussion

Children are at risk for sudden death if suffering from undiagnosed high-risk cardiac conditions. More needs to be done to prevent unnecessary and preventable deaths from sudden cardiac arrest. We may not have an Italian experience (3) of screening all athletes, or a Japanese experience (where all children are screened throughout their growing years into adolescence, but the children and families deserve to know the risks of sudden cardiac death and the means to mitigate such risks through affordable cardiac screenings. At the time of Anthony’s death, 20 years ago, there were only two organizations hosting free or low-cost (under $100 for youth, under $200 for adults) cardiac screenings for youth and their families. Currently, there are nearly 1,000 non-profit organizations and hospitals nationwide hosting events to prevent sudden cardiac arrest in the young.

By 2019, ABF had screened over 15,600 young hearts, finding thirteen percent with problems and a consistent two percent with “Possible Life-Threatening” issues. There are more than seventy-five active ABF Trained Teams in the U.S. Each team is hosting community heart screenings; 2020 figures showed the young hearts screened by our ABF Trained Teams to be more than 1,013,745. Our ABF Trained Teams are training other teams. Including these numbers, in our count, we have surpassed 1 million screened before 2020 – even surrounded by this COVID-19 pandemic (Bates, 2020, p. 164).

Heart screening for youth and families is a reality in our country. More can be done and will be done to protect the precious lives of our country’s youth using ultrasound with ECG tests to the validity of the findings. These tests are low cost, affordable, and should be available for all people in our country, not just athletes and not just one screen. The findings of repeat heart screens in this review of a 20-year experience of heart screens show the alarming rate of increase of abnormal findings from the overall ABF heart-screen database to 17.49%. There are both congenital cardiac anomalies and acquired cardiac anomalies that are among the abnormal heart-screen findings. More review, research, and data can be collected, but until heart screenings are given more priority in healthcare, these will only be done through the efforts of the grieving parents in our country. Through our screening, we reported numerous correlations between our screening and clinical presentations.

We could record accurate prevalence of mitral valve prolapse and suspected hypertrophic cardiomyopathy in a young population (6,7) , Furthermore we recorded correlation between obesity with hypertension, high heart rate and symptoms. (8–11). We also reported that normal EKG and absence of physical symptoms were found in substantial number of our participant with suspected hypertrophic cardiomyopathy (12,13) . We also documented race relation to abnormal EKGs (13, 14). Cost effectiveness of performing echocardiographic screening has also been well documented. In a screening of 2,111 athletes, 30 was found to have conditions associated with sudden cardiac death (14) with little cost by adding EKG to the screening with additional benefits (15,16). Cost effectiveness calculations clearly support screening athletes. (17). Hopefully, screening pick up steam based on our 20 years screening experience and our findings will help to developed criterion for more cost effective and comprehensive screening.

## Conclusions

Cardiac screening involving multiple repeated screenings appears to be effective in detecting increasing numbers of abnormal findings that can be lifesaving.

## Limitations

our study is a retrospective study and not a prospective controlled trial. Screening involved volunteers who may not represent general population.

## Data Availability

Data not publically available

## Notes

### Competing Interest Statement

The authors have declared no competing interest.

### Clinical Trial

N/A

### Funding Statement

No founding

### Author Declarations

It was approved and Exempt by University of Arizona IRB

## References

1. Maron BJ, Poliac LC, Roberts WO. Risk for sudden cardiac death associated with marathon running. J Am Coll Cardiol. 1996 Aug;28(2):428–31

2. Bates, S. Damage control - heart breaks to heart saves. Phoenix: Sharon Bates, Anthony Bates Foundation., Book, 2020, ISBN-13: 9781641844376

3. Link MS, Estes NA 3rd. Sudden cardiac death in the athlete: bridging the gaps between evidence, policy, and practice. Circulation. 2012 May 22;125(20):2511–6.

4. Kaltman JR, Thompson PD, Lantos J, Berul CI, Botkin J, Cohen JT, Cook NR, Corrado D, Drezner J, Frick KD, Goldman S, Hlatky M, Kannankeril PJ, Leslie L, Priori S, Saul JP, Shapiro-Mendoza CK, Siscovick D, Vetter VL, Boineau R, Burns KM, Friedman RA. Screening for sudden cardiac death in the young: report from a national heart, lung, and blood institute working group. Circulation. 2011:3;123(17):1911–8.

5. Graham R, McCoy MA, Schultz AM. Strategies to Improve Cardiac Arrest Survival: A Time to Act. Washington (DC): National Academies Press (US); 2015, https://www.ncbi.nlm.nih.gov/books/NBK305685/ doi: 10.17226/21723

6. Sattur S, Bates S, Movahed MR. Prevalence of mitral valve prolapse and associated valvular regurgitations in healthy teenagers undergoing screening echocardiography. Exp Clin Cardiol. 2010;15(1):e13–5.

7. Movahed MR, Strootman D, Bates S, Sattur S. Prevalence of suspected hypertrophic cardiomyopathy or left ventricular hypertrophy based on race and gender in teenagers using screening echocardiography. Cardiovasc Ultrasound. 2010: 10;8:54.

8. Movahed MR, Bates S, Strootman D, Sattur S. Obesity in adolescence is associated with left ventricular hypertrophy and hypertension. Echocardiography. 2011;28(2):150–3.

9. Hickcox L, Bates S, Movahed MR. Presence of physical symptoms in healthy adolescence found to be associated with female gender, obesity, tachycardia, diastolic hypertension and smoking. Am J Cardiovasc Dis. 2022;12(6):315–319.

10. Movahed MR, Motieian M, Bates S. Overweight (BMI of 25-30) Is Independently Associated With Significantly Higher Prevalence of Systolic and Diastolic Hypertension in Adults. Crit Pathw Cardiol. 2023;22(4):146–148.

11. Hickcox L, Bates S, Hashemzadeh M, Movahed MR. Higher Heart Rate Is Independently Associated With Abnormal Body Mass Index in a J Shape Pattern. Crit Pathw Cardiol. 2023;22(3):100–102

12. Movahed MR, Sattur S, Bates S. Higher Prevalence of Abnormal Electrocardigrams (ECG) in African Americans Undergoing Screening ECG and Echocardiography. Crit Pathw Cardiol. 2019:(2):86–88.

13. Movahed MR, Ramaraj R, Bates S. Early repolarization is independently associated with African-American race, younger age, higher BMI and lower heart rate. Future Cardiol. 2022;18(10):771–775

14. Halasz G, Capelli B, Nardecchia A, Cattaneo M, Cassina T, Biasini V, Barbieri D, Villa M, Beltrami M, Perone F, Villani M, Badini M, Gervasi F, Piepoli M, Via G. Cost-effectiveness and diagnostic accuracy of focused cardiac ultrasound in the pre-participation screening of athletes: the SPORT-FoCUS study. Eur J Prev Cardiol. 2023 ;30(16):1748–1757.

15. Wheeler MT, Heidenreich PA, Froelicher VF, Hlatky MA, Ashley EA. Cost-effectiveness of preparticipation screening for prevention of sudden cardiac death in young athletes. Ann Intern Med. 2010;152(5):276–86.

16. Baggish AL, Hutter AM Jr, Wang F, Yared K, Weiner RB, Kupperman E, Picard MH, Wood MJ. Cardiovascular screening in college athletes with and without electrocardiography: A cross-sectional study. Ann Intern Med. 2010;152(5):269–75.

17. Fuller CM. Cost effectiveness analysis of screening of high school athletes for risk of sudden cardiac death. Med Sci Sports Exerc. 2000;32(5):887–90

